## Supplementary Figure Legends for "“When will this end? Will it end?” The impact of the March-June 2020 UK Covid-19 lockdown response on mental health: a longitudinal survey of mothers in the Born in Bradford study"

**Supplementary Figure 1: PHQ-8 at pre-Covid19 and Covid19 lockdown survey with change between categories**

***Note: This graph presents findings from participants who had complete pre-Covid19 and Covid19 lockdown PHQ-8 scores (n=1730)***

**Supplementary Figure 2: GAD-7 at pre-Covid19 and Covid19 lockdown survey with change between categories**

***Note: This graph presents findings from participants who had complete pre-Covid19 and Covid19 lockdown GAD-7 scores (n=1634)***
