## Supplementary figures and images for "“When will this end? Will it end?” The impact of the March-June 2020 UK Covid-19 lockdown response on mental health: a longitudinal survey of mothers in the Born in Bradford study"

### Supplementary Figure 1

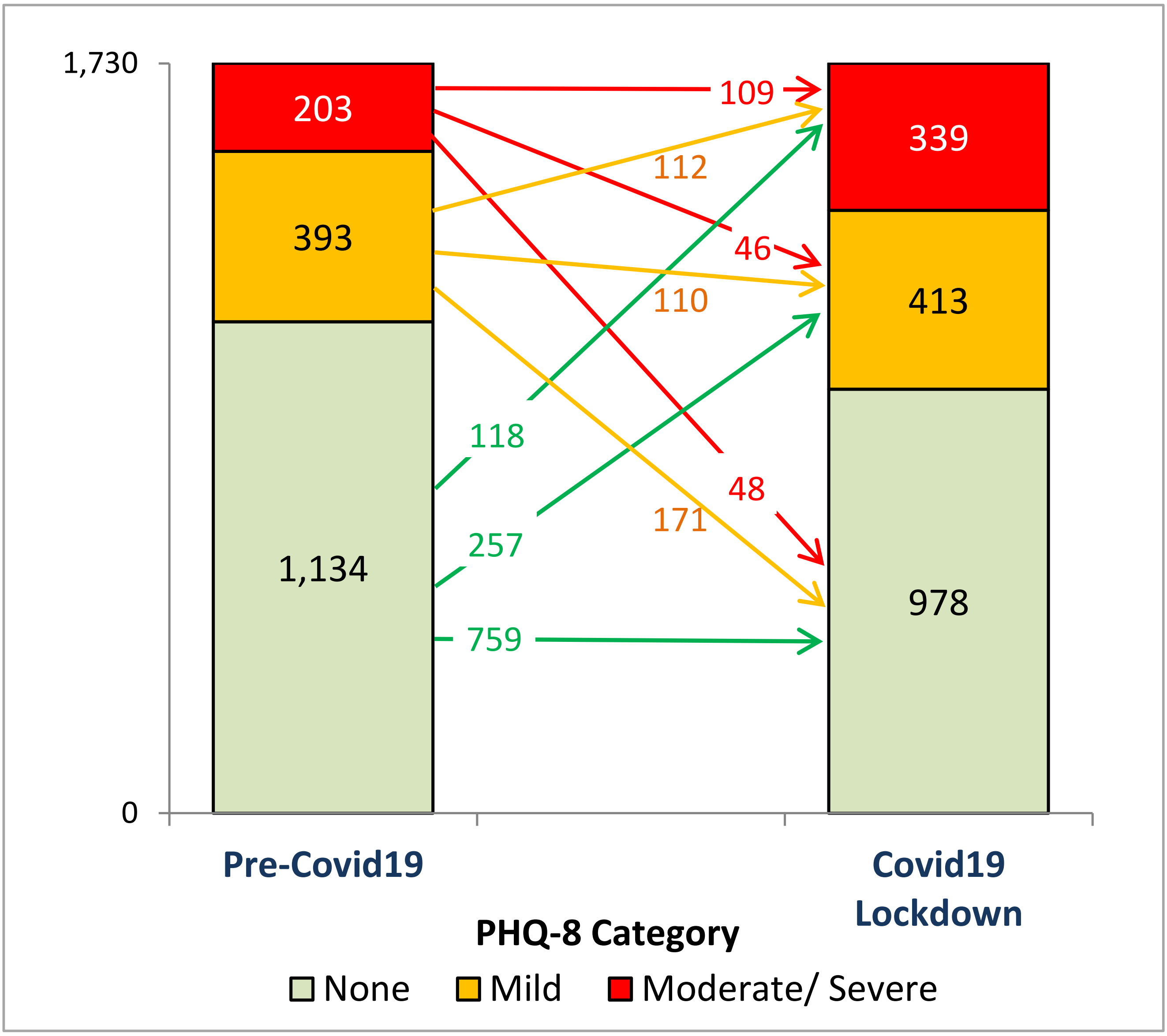

### Supplementary Figure 2

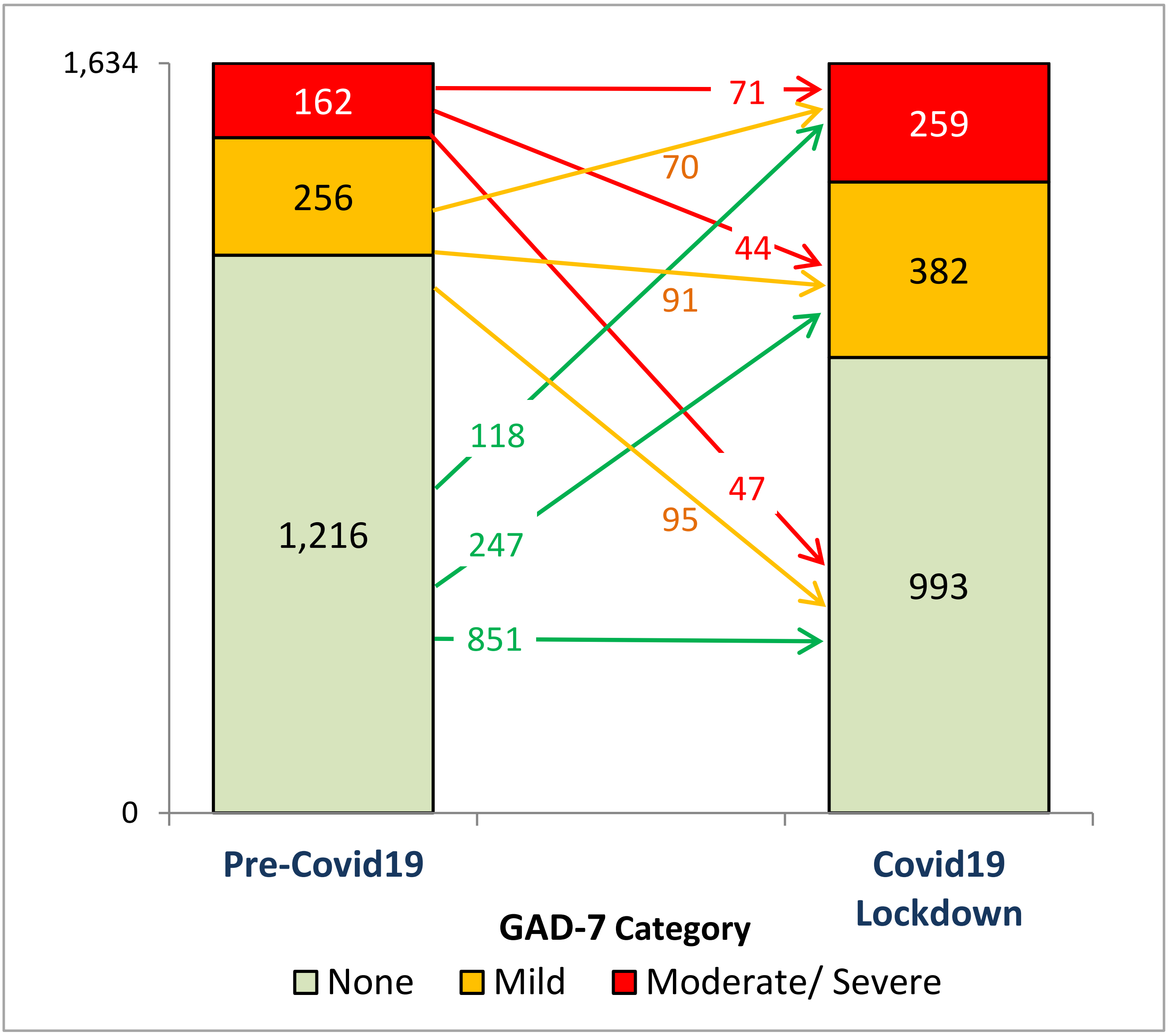
