## Supplementary Tables 1-2 for "“When will this end? Will it end?” The impact of the March-June 2020 UK Covid-19 lockdown response on mental health: a longitudinal survey of mothers in the Born in Bradford study"

Supplementary Table 1: The sample characteristics of those invited to complete the Covid-19 survey by survey completion status.

| | Returned Survey (n=2144) | | | |
|---|---|---|---|---|
| **Age** | Complete survey (n=1860) | Incomplete / not linked (n=284) | Not returned survey (n=4722) | Total eligible (n=6866) |
| Under 30 yrs. | 224 (12%) | 4 (2%) | 751 (16%) | 963 (14%) |
| 30 to 34 yrs. | 396 (21%) | 22 (10%) | 1182 (25%) | 1600 (24%) |
| 35 to 39 yrs. | 516 (28%) | 66 (29%) | 1367 (29%) | 1939 (29%) |
| 40 to 44 yrs. | 423 (23%) | 73 (32%) | 892 (19%) | 1395 (21%) |
| 45 yrs. plus | 301 (16%) | 61 (27%) | 495 (11%) | 887 (13%) |
| Missing | - | 58 | 35 | 82 |
| **Ethnicity*** | | | | |
| White British | 613 (34%) | 102 (47%) | 821 (18%) | 1527 (23%) |
| Pakistani Heritage | 877 (48%) | 80 (37%) | 2921 (64%) | 3843 (59%) |
| Other | 320 (18%) | 36 (17%) | 803 (18%) | 1148 (18%) |
| Missing | 50 | 66 | 177 | 348 |
| **Depression (PHQ8)** | | | | |
| None | 1187 (66%) | - | 2594 (64%) | 3774 (64%) |
| Mild | 414 (23%) | - | 945 (23%) | 1345 (23%) |
| Moderate | 135 (7%) | - | 346 (8%) | 475 (8%) |
| Moderately severe | 58 (3%) | - | 134 (3%) | 193 (3%) |
| Severe | 19 (1%) | - | 64 (2%) | 83 (1%) |
| Missing | 47 | 284 | 639 | 996 |
| **Anxiety (GAD7)** | | | | |
| None | 1280 (75%) | - | 2720 (72%) | 3991 (73%) |
| Mild | 270 (16%) | - | 646 (17%) | 908 (17%) |
| Moderate | 100 (6%) | - | 224 (6%) | 322 (6%) |
| Severe | 67 (4%) | - | 166 (4%) | 234 (4%) |
| Missing | 143 | 284 | 966 | 1,411 |

**Note: The ethnicity representativeness is skewed by the BiBBS participants who are ~70% South Asian and had a comparatively low response rate to this survey – see reference 15 for explanation*
